# Effectiveness of inactivated and Ad5-nCoV COVID-19 vaccines against SARS-CoV-2 Omicron BA. 2 variant infection, severe illness, and death

**DOI:** 10.1101/2022.09.04.22279587

**Authors:** Zhuoying Huang, Shuangfei Xu, Jiechen Liu, Linlin Wu, Jing Qiu, Nan Wang, Jia Ren, Zhi Li, Xiang Guo, Fangfang Tao, Jian Chen, Donglei Lu, Xiaodong Sun, Weibing Wang

**Author notes:** Corresponding: Dr. Xiaodong Sun, Shanghai Municipal Center of Disease Control and Prevention, 1380 Western Zhongshan Road, Shanghai 200336, China., Dr. Weibing Wang, Department of Epidemiology, School of Public Health, Fudan University, 138 Yi Xue Yuan Road, Shanghai 200032, China. These authors contributed equally to this work (Zhuoying Huang, Shuangfei Xu).

## Abstract

**Background:** Limited data are available on effectiveness of inactivated and Ad5-nCoV COVID-19 vaccines in real-world use - especially against Omicron variants in SARS-CoV-2 infection-naïve population. During an outbreak in Shanghai’s SARS-CoV-2 infection-naïve population, we evaluated vaccine effectiveness (VE) against Omicron infection, severe or critical COVID-19, and COVID-19-related death.

**Methods:** A matched case-control study was conducted among people aged ≥3 years between 2 December 2021 through 13 May 2022. Cases were SARS-CoV-2 infected individuals, individuals with severe/critical COVID-19, or COVID-19-related deaths. Controls were selected from consecutively test negative individuals at the same time as cases were diagnosed and were exact-matched on year-of-age, gender, birthplace, illness onset date, and residency district in ratios of 1:1 with infected individuals and 4:1 with severe/critical COVID-19 and COVID-19-related deaths.

**Results:** Our study included 612597 documented SARS-CoV-2 infections, among which 1485 progressed to severe or critical illness and 568 died. Inactivated vaccine was 16.3% (95% CI: 15.4%-17.2%) effective against infection, 88.6% (95% CI: 85.8%-90.9%) effective against severe/critical COVIID-19 and 91.7% (95% CI: 86.9%-94.7%) against COVID-19 death. Ad5-vectored vaccine was 13.2% (95% CI: 10.9%-15.5%) effective against infection and 77.9% (95% CI: 15.6%-94.2%) effective against severe/critical COVIID-19. Booster vaccination with inactivated vaccines enhanced protection against severe COVID-19 (92.7%, 95% CI: 90.1%-94.6%) and COVID-19 death (95.9%, 95% CI: 91.4%-98.1%). Inactivated VE against infection began to wane 12 weeks after the last dose but two- and three-dose sustained high protection levels (>80%) against severe/critical illness and death.

**Conclusions:** Our real-world study found high and durable two- and three-dose inactivated VE against Omicron-associated severe/critical illness and death across all age groups, but lower effectiveness against Omicron infection. High direct protection from severe/fatal Omicron COVID-19 provided by inactivated vaccines, and a consequent potential reduction in health-care utilization, reinforces the critical importance of full-series vaccination and timely booster dose administration for all eligible individuals.

## INTRODUCTION

The Omicron BA. 2 SARS-CoV-2 variant of concern was first detected in the United States in November of 2021 and has subsequently spread globally. ^1^ Sequencing of virus isolates from 129 COVID-19 patients between late February 2022 and May 2022 showed that Omicron BA. 2 was the dominate sub-lineage in Shanghai^2^. Omicron BA. 2 differs by approximately 40 mutations from the original Omicron lineage, BA. 1, and has a growth advantage over BA.1. ^3^ While studies are ongoing to understand the reasons for this growth advantage, initial data suggest that BA.2 appears more transmissible than BA. 1, which currently remains the most common Omicron sub-lineage reported. Rodent models suggest that infectivity and pathogenicity of Omicron/BA. 2 variant are similar to Omicron/BA. 1’s.^4,5^

Studies in Israel, the United Kingdom, and the United States have demonstrated that primary-series and booster vaccination with mRNA vaccines protect against SARS-CoV-2 infection, with greater protection from COVID-19 related hospitalizations and severe outcomes ^6-8^ which is also seen against Omicron outcomes. ^9-11^ Studies have also reported reduced neutralizing antibody activity against Omicron. ^12^ Fewer data are available on real-world vaccine effectiveness (VE) of inactivated and adenovirus type 5 vectored vaccine, especially against Omicron. A Singapore study before Omicron became prevalent reported that compared with BNT162b2, subjects who received inactivated whole virus vaccines were more likely to be infected with SARS-CoV-2. ^13^ An ecological VE study in Hong Kong found that three doses of either BNT162b2 or CoronaVac provided substantial protection against severe COVID-19 (VE>95%) and death (VE>96%) caused by confirmed Omicron variant infection. ^14^

In Shanghai, COVID-19 vaccination started in February 2021 among residents 18-59 years old, in March 2021 among residents 60-75 years of age, in May 2021 for individuals 76 years and older, and in September 2021 for children 3-17 years of age. Booster vaccination started in November 2021 for residents ≥ 18 years; children are not yet eligible for boosters. As of May 2022, there is no approved COVID-19 vaccine for children <3 years in China. One out of every four doses of COVID-19 administered globally has been CoronaVac. Despite its global importance, limited evidence is available on the efficacy or effectiveness of this vaccine.

Between 2 December 2021 and 13 May 2022, a total of 618,019 individuals tested RT-PCR-positive for SARS-CoV-2 infection. By that time, over 90% of the population aged ≥ 3 years were reported to have received a primary series, and 47% received a booster dose. With its SARS-CoV-2-infection-naïve population, Shanghai provided a nearly-unique outbreak setting to estimate real-world, vaccine-only-induced inactivated and Ad5-vectored COVID-19 VE against documented SARS-CoV-2 Omicron infection, severe or critical COVID-19, and COVID-19 related death.

## METHODS

### Study Setting and Study Participants

The study was conducted among people living in Shanghai, a provincial-level municipality in China with a resident population of more than 25 million people. Everyone living in Shanghai - citizens, foreigners, and immigrants - underwent several rounds of SARS-CoV-2 real time polymerase chain reaction (RT-PCR) testing between 2 December 2021 and 13 May 2022. Complete demographic (age, gender, birthplace, and residency district), vaccination, and RT-PCR testing data were available to public health officials and study investigators. Individuals with negative RT-PCR SARS-CoV-2 tests who had tested positive before 2 December 2021 were excluded. Children less than three years of age were not vaccine-eligible and were excluded.

Controls were selected from consecutively test negative individuals at the same time as cases were diagnosed. The size of the potential control group was 25.18 million people receiving several round city-wide nucleic acid amplification testing (NAAT), and RT-PCR testing was universal regardless of COVID-19 associated symptoms.

### Study Design

We used a matched case-control design for data extraction. Cases were individuals with documented SARS-CoV-2 infection, severe or critical COVID-19, or COVID-19-related death. Infection-only cases were matched 1:1 with controls on year-of-age, gender, birthplace (Shanghai/other place), date of testing, and residency district; severe or critical COVID-19 cases and COVID-19-related death cases were matched 1:4 on year-of-age, gender, birthplace, date of illness onset, and residency district (Figure 1). We matched each case to unique controls without replacement. When there are more than one case-control pair available, we selected the first (for infection-only cases) or first four (for severe/critical and fatal cases) eligible individuals. We constructed two subsets to analyze separate VEs of inactivated vaccines (subset 1) and Ad5 vectored vaccine (subset 2) against each of the three outcomes (figures S1 and S2).

**Figure 1.**
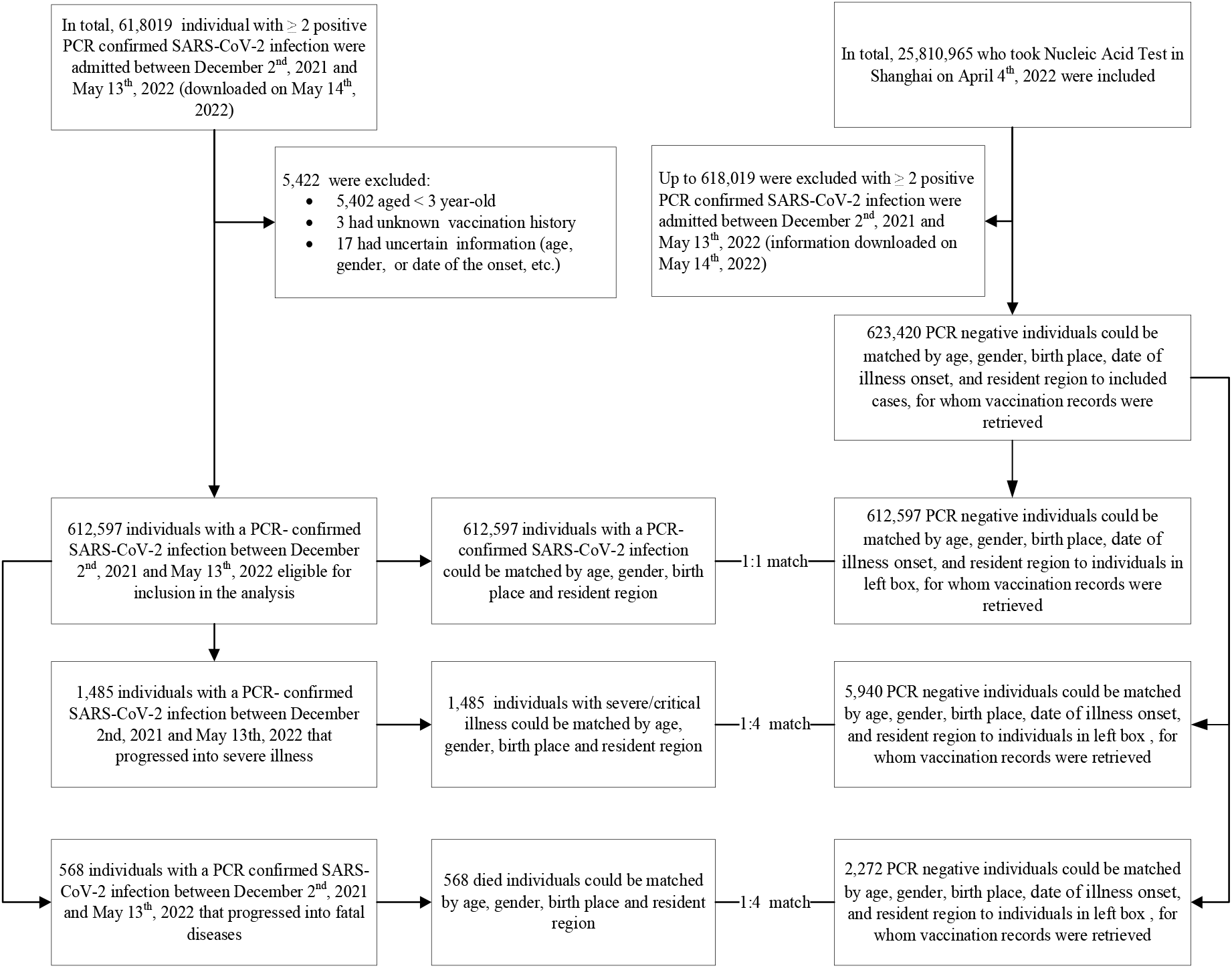
Participant enrollment flowchart.

### Infections and Outcomes

PCR testing for SARS-CoV-2 is available in public hospitals and private laboratories throughout China. Since 10 March 2022, Shanghai conducted several rounds of SARS-CoV-2 tests, each involving more than 25 million people. Severity of disease for all SARS-CoV-2 infections was assessed in any of 48 COVID-19 designated hospitals in accordance with the *Diagnosis and Treatment Protocol for COVID-19 (Trial Version 9)*. ^15^ We included three outcomes in our study: documented SARS-CoV-2 infection, severe/critical COVID-19, and COVID-19-related death. Documented SARS-CoV-2 infection was confirmed by a positive RT-PCR test. For adult cases, severe illness must meet any of the following criteria: a) respiratory distress (Respiration Rate [RR] ≥ 30 breaths per min), b) oxygen saturation ≤ 93% at rest, c) arterial partial pressure of oxygen/fraction of inspired oxygen ≤ 300mmHg. Additionally, cases with chest imaging that shows obvious lesion progression within 24-48 hours > 50% shall be managed as severe illness. For child cases, severe illness must meet any of the following criteria: a) Persistent high fever over three days, b) tachypnea (RR ≥ 60 BPM for infants aged below 2 months; RR ≥ 50 BPM for infants aged 2-12 months; RR ≥ 40 BPM for children aged 1-5 years, and RR ≥ 30 BPM for children >5 years), independent of fever and crying, c) oxygen saturation ≤93% on finger pulse oximeter taken at rest, d) labored breathing (moaning, nasal fluttering, infrasternal, supraclavicular and intercostal retraction), cyanosis, and intermittent apnea, e) lethargy and convulsion, f) difficulty feeding and signs of dehydration. Critical illness must meet any of the following criteria: a) respiratory failure and requiring mechanical ventilation; b) shock, c) With other organ failure that requires ICU care. COVID-19 related death is assessed by medical institutions.

Data on all confirmed and asymptomatic cases between 2 December 2021 and 13 May 2022 were extracted from the National Notifiable Diseases Registry System (NNDRS).

### Vaccination Data

The Shanghai Group Immunization System captures all vaccine administrations and is updated daily. This immunization information system is the repository of nearly all vaccinees’ records, and includes name, national identification number, vaccine type, vaccination date, vaccination dose, vaccination site, and vaccine manufacturer. This system is linked to the National Immunization Program Information System, which adds documented national identification-matching COVID-19 vaccinations received outside of Shanghai. Immunization histories were verified manually with records for subjects three years and older who had an unknown national identification number. NAAT data were linked to individual vaccination records using national identification number and name. Vaccination data were extracted and matched to cases and controls on 13 May 2022. Figure S3 shows the process of vaccination history retrieval.

Vaccination status was categorized into four levels in accordance with national technical recommendations for COVID-19 vaccination: (1) unvaccinated – either no history of COVID-19 vaccination before the last SARS-CoV-2 exposure date; (2) partial vaccination – either one dose of inactivated vaccine; or two doses of inactivated vaccine but receiving the second dose within 14 days before the last SARS-CoV-2 exposure date; two doses of recombinant protein vaccine (three doses are recommended for primary vaccination); or one dose of adenovirus vector vaccine or three doses of recombinant protein vaccine but with the most recent dose within 14 days before the last SARS-CoV-2 exposure date; (3) full primary vaccination – either two doses of inactivated vaccine, one dose of adenovirus vector vaccine, or three doses of recombinant protein (CHO cell) vaccine with the most recent doses 14 days or more before the last SARS-CoV-2 exposure date and with no booster dose; or two doses of inactivated vaccine with one booster dose of inactivated vaccine, adenovirus vector vaccine, or recombinant protein (CHO cell) vaccine within 7 days before the last SARS-CoV-2 exposure date; or two doses of adenovirus vector vaccine within 7 days before the last exposure date; or (4) booster vaccination – either two doses of Ad5-vectored vaccine with the second dose 7 days or more before the last exposure date; or two doses of inactivated vaccine and one booster dose of inactivated vaccine, adenovirus vector vaccine, or recombinant protein vaccine 7 days or more before the last exposure date.

Vaccination intervals were stratified by dose and interval (in weeks) post vaccination at <2, 3-12, 13-24, 25-36, 37+ weeks after the first dose, <2, 3-12, 13-24, 25-36, 37+ weeks after the second dose, and <2, 3-12, 13-24, 25+ weeks after the third dose. Intervals (in days) were defined as the number of days between the onset date of infection and the date of last vaccination minus 2 days, which was then converted into weeks. Detailed variable definitions are in table S1. Additionally, we performed a sensitivity analysis in which intervals (in days) were defined as the number of days between testing positive date and the date of last vaccination minus 7 days.

### Statistical Analysis

Non-normally distributed continuous variables were expressed as medians (interquartile ranges, IQR), and categorical variables were expressed as counts and proportions. Conditional logistic regression was used to estimate the odds ratio (OR) of vaccination among cases and controls, with documented SARS-CoV-2 infection, severe or critical COVID-19, and COVID-19 related death as dependent variables. Vaccination status was the independent variable and VE was defined as 1 minus the matched OR. Vaccination status was defined using the date of onset and date of last vaccination, as above. Analyses were stratified by age category, gender, and vaccine type (inactivated vaccine, adenovirus vector vaccine, and recombinant protein vaccine). Matching were conducted with the use of JAVA, and Analyses were performed with the use of RStudio 2022.02.3+492.

## RESULTS

### Study Population

There were 612597 SARS-CoV-2 RT-PCR confirmed cases in the study, among which 1485 progressed into severe or critical illness and 568 died (Table 1). The most common age for infection was 40-59 years of age (37.3%); the most common age for severe/critical COVID-19 and COVID-19 death was 80 years and older (58.6% of severe/critical COVID-19 and 69.9% of COVID-19 deaths). Due to the exact matching, the demographic characteristics of controls and cases were the same (Table 1).

### Vaccination Status

Figure 2 shows vaccination status of the control group over time and by age group. At the beginning of the study period, 82.7% people received at least one dose of a COVID-19 vaccine, with the majority 18-59-years old. The prevalence of the adult-focused booster dose increased from 11.9% to 49.0%. Full and partial coverage among adults aged 80 years and above was low and stable at approximately 15%. Figure 2 also shows the cumulative incidence of documented SARS-CoV-2 infection (250.9 per 10,000), of which the majority were 18-59 years old. Since March 12, a series of emergency epidemic prevention measures were implemented in Shanghai, including mass NAATs, citywide lockdowns, home quarantine of residents, isolation of cases, centralized quarantine of close contacts, and closed-loop management. The rapid spread of Omicron led to temporary suspension of vaccination starting in March 2022.

**Figure 2.**
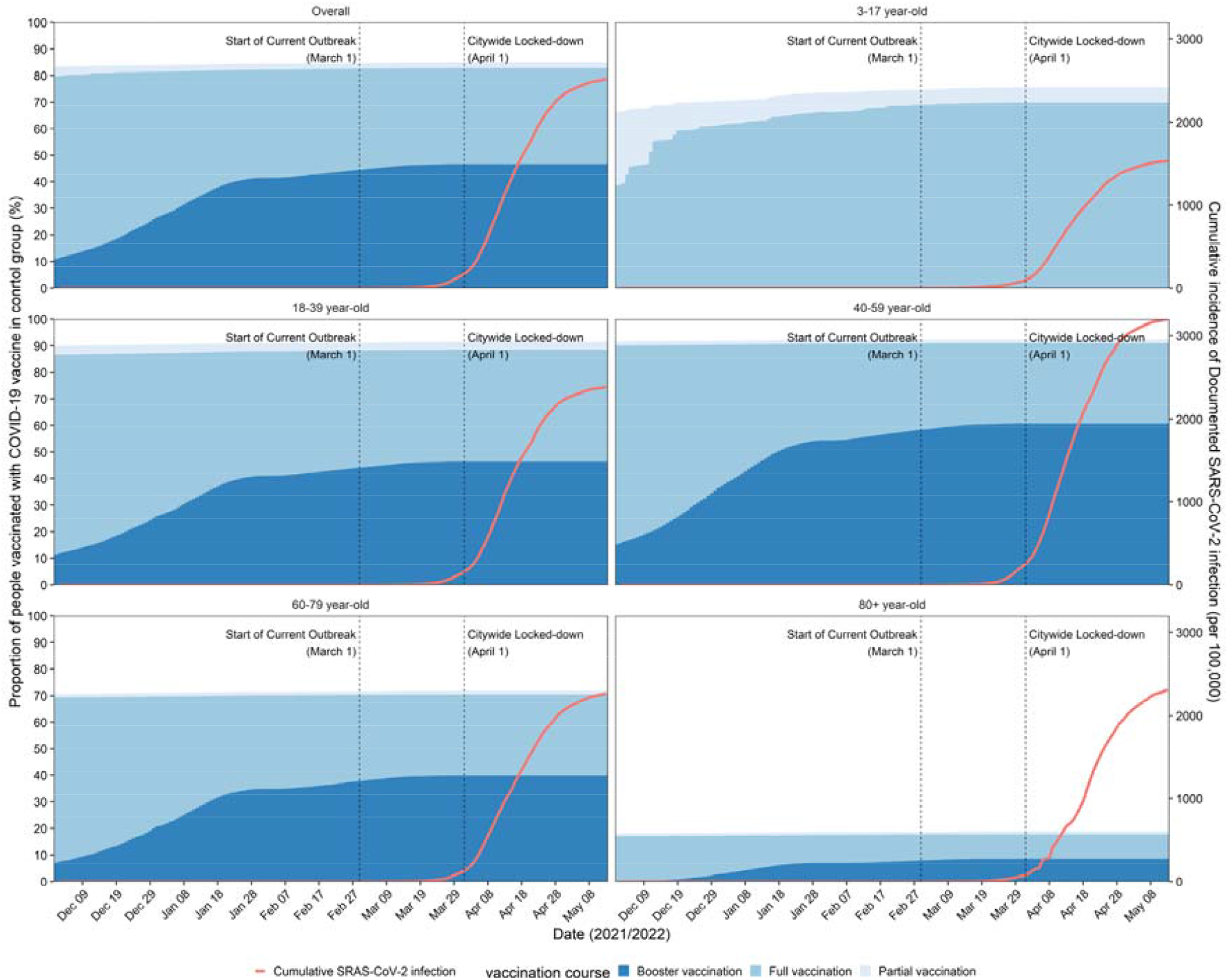
Vaccination status in the control group and cumulative documented SARS-CoV-2 infections by age grouping in Shanghai, 1 December 2021 through 13 May 2022. Also shown are public health and social measures implemented during the study period. Vaccination status for control group was from a total of 623,420 controls, including 1:1 match with infected individuals and 4:1 with severe/critical COVID-19 and COVID-19-related deaths. Data for cumulative incidence of SARS-CoV-2 infection was provided by Shanghai Health Commission, and numerator was the daily number of cumulative infected cases and denominator was the corresponding population size from the seventh National Census.

### Vaccine Effectiveness

COVID-19 vaccines provided some protection against SARS-CoV-2 infection in all age groups (overall VE: 16.0%, 95% CI: 15.1%, 17.0%), while in age group 40-59 years, protection was lower (VE: 7.5%, 95%CI: 5.5%, 9.5%). COVID-19 vaccines provided high effectiveness against severe/critical illness (VE: 88.6%, 95% CI: 85.8%, 90.8%) and COVID-19 related death (VE: 91.6%, 95% CI: 86.8%, 94.6%). There was little difference in VE by gender. Estimated VEs for full primary vaccination and booster doses against severe/critical illness were 83.8% (95% CI: 79.0%, 87.5%) and 92.8% (95% CI: 90.2%, 94.7%), respectively, and against COVID-19 related death were 85.7% (95% CI: 75.6%, 91.6%) and 96.2% (95% CI: 92.0%, 98.2%), respectively. Inactivated vaccine was 16.3% (95% CI: 15.4%, 17.2%) effective against infection, 88.6% (95% CI: 85.8%, 90.9%) effective against severe/critical COVIID-19 and 91.7% (95% CI: 86.95%, 94.7%) against COVID-19 death. Ad5-vectored vaccine was 13.2% (95% CI: 11.0%, 15.5%) effective against infection and 77.9% (95% CI: 15.6%, 94.2%) effective against severe/critical COVIID-19. See Table 2.

We used subset 1 and subset 2 to estimate vaccine specific VEs. Figure 3 shows VEs of inactivated vaccine against SARS-CoV-2 infection, severe/critical illness, and COVID-19 related death; respective overall VEs were 16.6% (95% CI: 15.6%, 17.5%), 88.5% (95% CI:85.6%, 90.7%) and 91.7 (95% CI: 86.8%, 94.8%). Booster vaccination with inactivated vaccine enhanced the protection for both severe outcome (VE: 92.7%, 95% CI: 90.1%, 94.6%) and death (VE: 95.9%, 95% CI: 91.4%, 98.1%).

**Figure 3.**
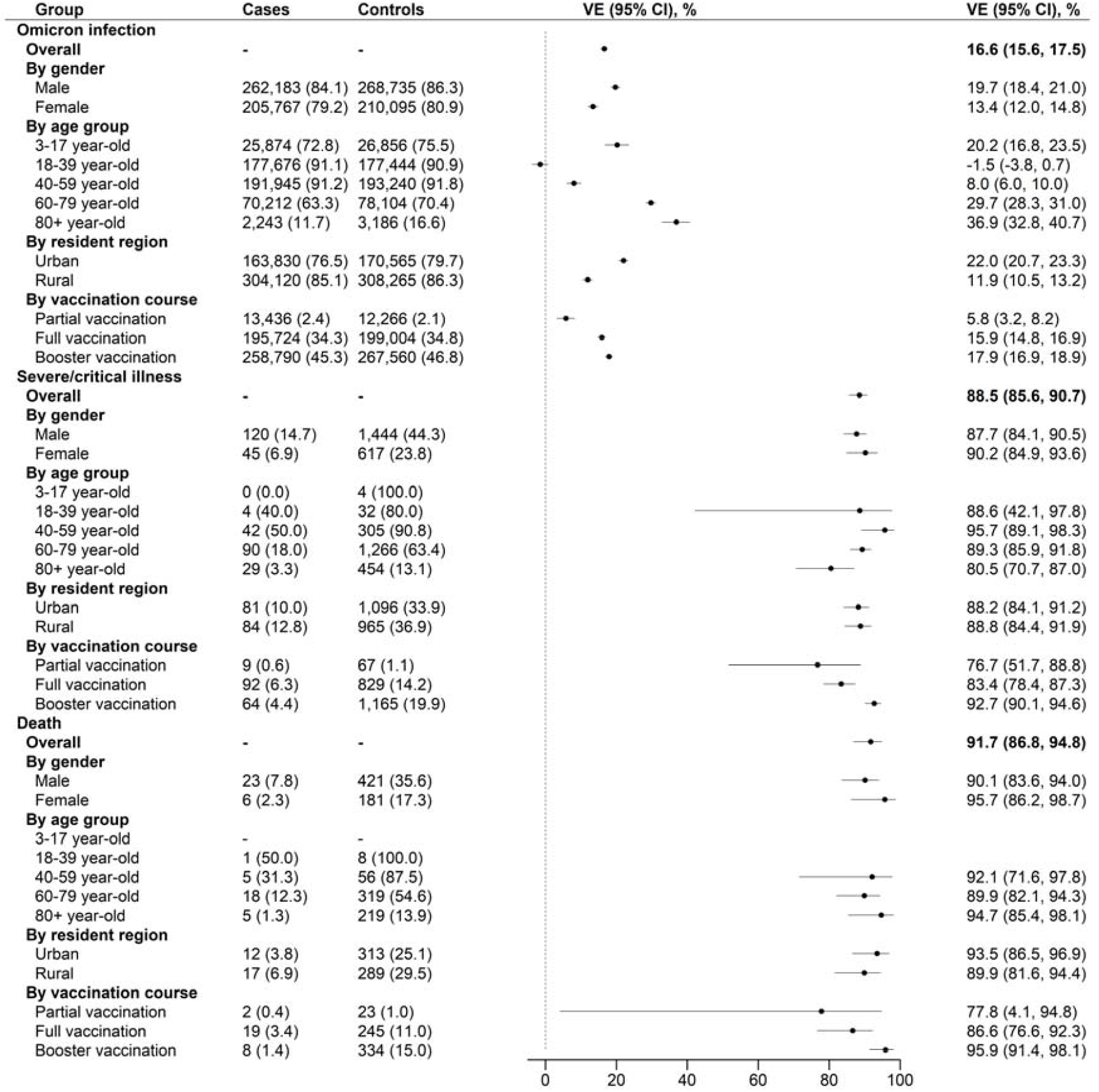
Vaccine effectiveness of inactivated vaccines against documented SARS-CoV-2 infection, severe or critical illness, and COVID-19 death in the matched case-control study.

Considering time between the last dose and SARS-Co2 exposure, figure 4 shows that the VE of inactivated vaccines against SARS-CoV-2 infection was 15.7% (95% CI:10.8%, 20.4%) 3-12 weeks after first dose. Two-dose inactivated vaccine offered 16.9% (95% CI: 13.2%, 20.4%) protection for infection after 12 weeks, and three-dose offered 19.2% (95% CI: 18.2%, 20.3%) after 24 weeks, then both subsequently becoming lower. Full series and boosted VEs for severe/critical illness and COVID-19 death remained >80%.

**Figure 4.**
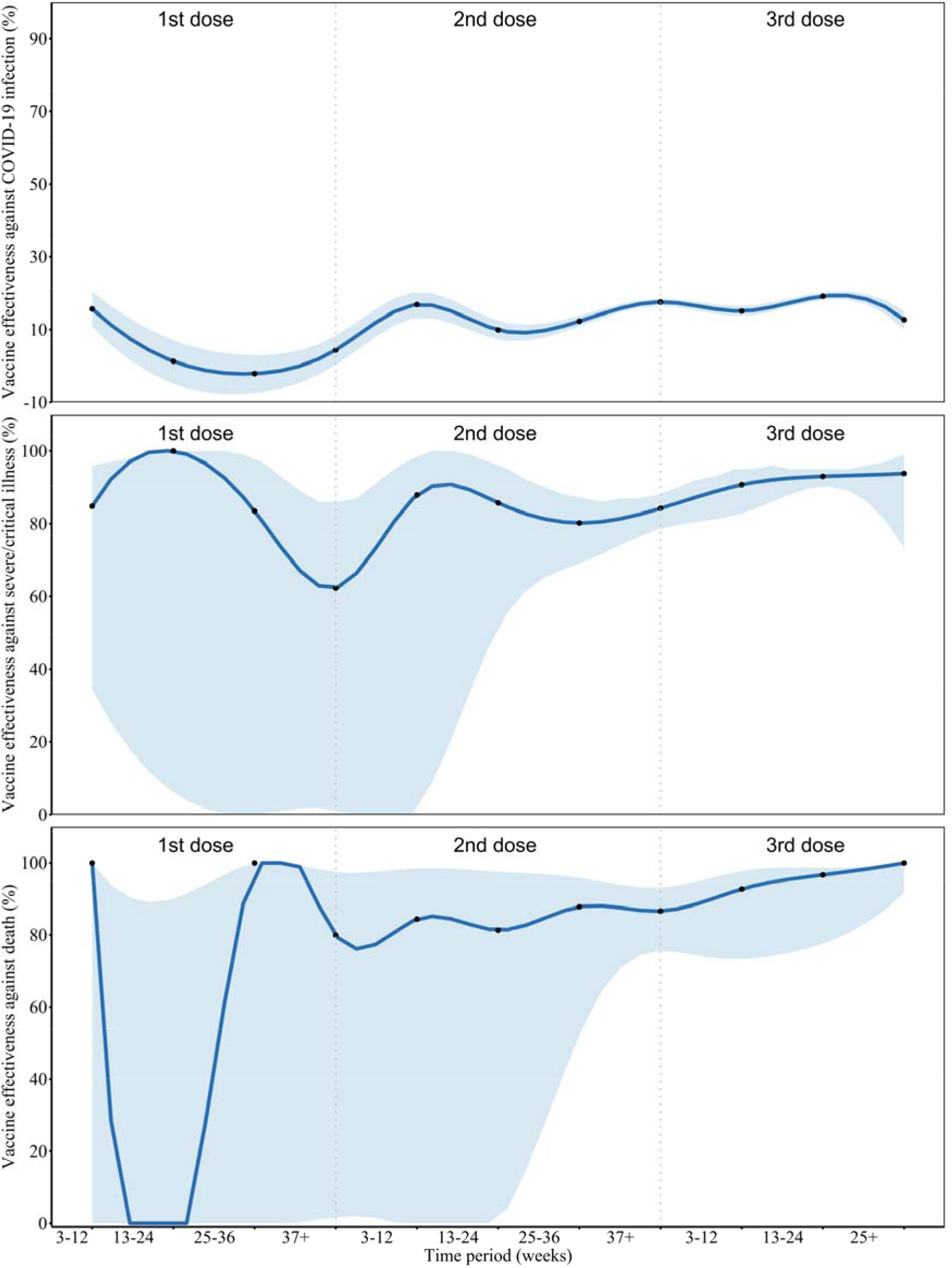
VE (%) of inactivated vaccine against documented SARS-CoV-2 infection, severe or critical illness, and death by doses and time interval. Data are presented with VEs (black points) and 95% confidence intervals (color zones). To visualize the trend of vaccine effectiveness (dark blue lines), we smoothed the VE results by performing three spline interpolations of the calculated data points for the corresponding time period. Small sample sizes of corresponding cells lead to the wide confidence intervals. Eight individuals had severe/critical illness and two died within first-doses intervals.

For SARS-CoV-2 infection, estimated VEs of Ad5-vectored vaccine were 39.3% (95% CI: - 17.9%, 68.8%) at 3-12 weeks after a first dose and 16.0% (95% CI: 1.5%, 28.4%) at 13-24weeks after a second dose. Due to the small number of severe/critical illness and death cases in subset 2, VEs by time interval were not able to be estimated for this vaccine (Table S2).

### Sensitivity Analysis

Tables S3 and S4 show the sensitivity analyses of VEs which were consistent with results displayed in table 2 and figure 3.

## DISCUSSION

From 2 December 2021 through 13 May 2022, during an Omicron BA2.2-predominant wave of COVID-19 in Shanghai, China, we used detailed, individual-level, records-documented COVID-19 vaccination status; universal, population-wide individual-level SARS-CoV-2 RT-PCR testing results; and individual-level clinical evaluations to estimate real-world effectiveness of inactivated and Ad5-vectored vaccines in a largely infection-naïve population. Our matched case-control study showed that the inactivated vaccines were highly effective against severe or critical COVID-19 and COVID-19-related death, findings that underscore the potential of the vaccines to save lives and substantially reduce demands on healthcare system. Inactivated vaccine was 16.3% effective against Omicron infection, 88.6% effective against severe/critical COVIID-19 and 91.7% against COVID-19 death. Ad5-vectored vaccine was 13.2% effective against infection and 77.9% effective against severe/critical COVIID-19. Booster-dose protection against severe/critical COVID-19 was 92.7% and against COVID-19 death was 95.9%. Protection from infection waned after 12 weeks, but primary series and booster-dose protection was sustained above 80% for at least 36 weeks from the last dose.

We found that VE against Omicron infection was lower than efficacy against ancestral-strain SARS-CoV-2 symptomatic infection observed in a phase 3 clinical trial (72.8% and 78.1%) and a real-world, pre-Omicron VE study in Chile (83.5%). ^16,17^ However, our VE estimates for the two predominant inactivated (Sinovac and Sinopharm) vaccines were very high in age-group analyses, notably among persons 80 years of age or older, and independent of time since vaccination. The lower VE against Omicron infection in younger adults may be partly due to a higher force of infection in working-age adults or differences in willing to be vaccinated (although vaccination rates were very high in all ages under 60 years). ^18^

Booster vaccination was 93% and 96% effective against severe illness and death, somewhat higher than full primary series vaccination VEs. We found that inactivated VE against severe illness and COVID-19 associated death to be higher than respective VEs observed in Brazil (77.6% and 83.9%) ^19,20^ and Chile (90.3% and 86.3%)^17^, in which the circulating SARS-CoV-2 strains predated Omicron emergence. Clinical trials have shown inactivated vaccine efficacies to be lower than that of mRNA vaccines, but there are fewer real-world data on two- and three-dose inactivated VE against Omicron infection or hospitalization. ^16,21^ The Hong Kong ecological study found an initial VE from two doses of inactivated COVID-19 vaccine against Omicron infection to be 17.9% with rapid waning - similar to our results. ^14^ Also similar to our findings, the Hong Kong scientists showed inactivated VE against severe illness and death to be very high – 74% for primary vaccination and 98% for boosted vaccination. Our study extended the time frame for duration of inactivated VE, showing sustained protection against severe COVID-19 and COVID-19 death through 36 weeks.

Omicron BA. 2 COVID-19 disease is less severe than Delta and BA. 1 associated COVID-19 disease^22^, resulting in a lower risk of hospitalization and severe illness. Indeed, over 85% of the SARS-CoV-2 infections reported from Shanghai by China’s National Health Commission were asymptomatic (http://www.nhc.gov.cn/xcs/xxgzbd/gzbd_index.shtml). Milder infections may reflect increased replication of Omicron in the upper versus lower respiratory tract – a phenomenon that could contribute to more efficient transmission, leading to a higher absolute number of hospitalizations. Omicron infection enhances preexisting immunity elicited by vaccines but, on its own, may not confer broad protection against non-Omicron variants in unvaccinated individuals^23^. A study demonstrated that inactivated vaccines elicited SARS-CoV-2-specific memory T/B cell immunity, which are important for robust recall of protective responses against viral replication, and therefore induce longer-term protection against severe illness and deaths. ^24,25^ However, Omicron’s immune evasion, including evasion of mucosal immunity allows Omicron to escape existing antibodies and increase risk of reinfection, leading to a surge in infections. ^26,27^ Evaluations of vaccines that have potential to block infection/transmission (*e*.*g*., the inhaled Ad5-vectored vaccine by CanSino that may provide mucosal immunity) are especially important. ^28,29^

We estimated the VEs of Ad5-vectored vaccine to be 39.3% at 3-12 weeks after a first dose but 16.0% at 13-24 weeks after a second dose. A study in Brazil reported that whereas waning of VE against symptomatic COVID-19 was observed ≥90 days after homologous and heterologous boosters, waning against severe COVID-19 was only observed after a homologous booster, suggesting a need for adjustment of vaccination strategies - *e*.*g*. heterologous boosters for those who have received 2 doses of inactivated vaccines as their primary series. ^20^ None of the current COVID-19 vaccines block infection/transmission; our results also show that the vaccines we evaluated cannot make a permanent barrier to contain the Omicron transmission in community, despite high vaccine coverage.

Our study has strengths. The setting was a COVID-19 outbreak in the large and diverse socioeconomic population of Shanghai in which 2.7% of the population was infected by the Omicron BA. 2 variant of SARS-CoV-2 before the outbreak was stopped by public health and social interventions. In addition, because COVID-19 is being managed as a Level-1 infectious disease, all infected individuals are compulsively reported to the disease prevention and control system daily. Infections are always confirmed with additional RT-PCR testing, making diagnostic misclassification highly unlikely. The cases in our study were from a community of 25 million people, repeatedly screened for infection using RT-PCR. We used individual level exact matching to control for potential confounding factors of age, sex, residential district, birthplace, and illness onset time. Finally, our study measured VE levels in an infection-naïve population, allowing assessment of nearly pure vaccine-induced, real-world VE, avoiding distortion from hybrid immunity.

Limitations of our study include possible misclassification of disease status, which was judged by the COVID-19 designated hospitals. The definitions of severe/critical COVID-19 are objective, the misclassification of severe illness is unlikely because disease severity for all SARS-CoV-2 infection is made in accordance with the *Diagnosis and Treatment Protocol for COVID-19*. A second limitation is the lack of information on comorbidities, precluding analyses by comorbidity. We attempted to reduce potential bias by matching on sociodemographic characteristics. In addition, the younger-aged population is at highest risk of infection and may also have greater willingness or opportunity to get vaccinated, possibly leading to an underestimate VE among working-age adults. Third, the study was conducted during a strict lockdown - especially after April 4 - which may have biased the estimation of VE (*e*.*g*. more likely to have null hypothesis) by decreasing the force of infection (attack rate among susceptibles). However, the universal NAAT screenings and exact matching that included district helped ensure similar exposure risk of cases and controls and our sensitivity analysis showed no meaningful differences in results. Finally, the number of Ad5-vectorred vaccine doses used in Shanghai was too small for subgroup analyses and for estimating VE against death.

## CONCLUSION

Our study showed high and durable two- and three-dose inactivated VE against severe/critical COVID-19 and death caused by Omicron infection across all age group, but with lower effectiveness against Omicron infection. Everyone eligible should be vaccinated; individuals eligible for booster doses should receive timely boosters; improving coverage among the elderly is especially important. The vaccines being used in China are highly effective where it matters most - preventing severe illness and death – and are most effective when boosted.

## Data Availability

The data that support the findings of this study are available from the Z.H. and X.S., upon reasonable request.

## List of abbreviations

VE: vaccine effectiveness
CI: Confidence interval
RT-PCR: real time polymerase chain reaction
NAAT: nucleic acid amplification testing
RR: Respiration Rate
NNDRS: National Notifiable Diseases Registry System
IQR: interquartile ranges
OR: odds ratio

## Ethics approval and consent to participate

The study was approved by the Ethical Review Committee in the Shanghai Center for Disease Control and Prevention (approval number: 2022-20).

## Consent for publication

Not applicable.

## Author contributions

Z.H. conceptualized the study, collected data and drafted the manuscript. S.X. performed analysis and drafted the manuscript. J.L. performed data clean and controlled quality. L.W. assisted with study design. J.Q., N.W., J.R., Z.L., F.T., and D.L. collected data. X.G., and J.C. reviewed manuscript. X.S. conceptualized the study, assisted with the analysis, and reviewed and edited the manuscript. W.W. assisted with the analysis and reviewed and edited the manuscript.

## Acknowledgement

We sincerely appreciate all the valuable comments and suggestions from Dr. Lance Rodewald, Dr. Zundong Yin, Dr. Zhijie An, Dr. Huaqing Wang, Dr. Wenzhou Yu, and Dr. Lin Tang from Chinese Center for Disease Control and Prevention, which allowed us to greatly improve the quality of the manuscript.

